# Oral SARS-CoV-2 host responses predict the early COVID-19 disease course

**DOI:** 10.1101/2023.03.06.23286853

**Authors:** William T Seaman, Olive Keener, Wenwen Mei, Katie R Mollan, Corbin D Jones, Audrey Pettifor, Natalie M Bowman, UNC OBSc Working Group, Frank Wang, Jennifer Webster-Cyriaque

## Abstract

**Objectives:** Oral fluids provide ready detection of Severe acute respiratory syndrome coronavirus 2 (SARS-CoV-2) and host responses. This study sought to determine relationships between oral virus, oral anti-SARS-CoV-2-specific antibodies, and symptoms.

**Methods:** Saliva/throat wash (saliva/TW) were collected from asymptomatic and symptomatic, nasopharyngeal (NP) SARS-CoV-2 RT-qPCR+, subjects (n=47). SARS-CoV-2 RT-qPCR, N-antigen detection by immunoblot and lateral flow assay (LFA) were performed. RT-qPCR targeting viral subgenomic RNA (sgRNA) was sequence confirmed. SARS-CoV-2-anti-S protein RBD LFA assessed IgM and IgG responses. Structural analysis identified host salivary molecules analogous to SARS-CoV-2-N-antigen. Statistical analyses were performed.

**Results:** At baseline, LFA-detected N-antigen was immunoblot-confirmed in 82% of TW. However, only 3/17 were saliva/TW qPCR+. Sixty percent of saliva and 83% of TW demonstrated persistent N-antigen at 4 weeks. N-antigen LFA signal in three negative subjects suggested potential cross-detection of 4 structurally analogous salivary RNA binding proteins (alignment 19-29aa, RMSD 1-1.5 Angstroms). At entry, symptomatic subjects demonstrated replication-associated sgRNA junctions, were IgG+ (94%/100% in saliva/TW), and IgM+ (75%/63%). At 4 weeks, SARS-CoV-2 IgG (100%/83%) and IgM (80%/67%) persisted. Oral IgG correlated 100% with NP+PCR status. Cough and fatigue severity (p=0.0008 and 0.016), and presence of nausea, weakness, and composite upper respiratory symptoms (p=0.005, 0.037 and 0.017) were negatively associated with oral IgM. Female oral IgM levels were higher than male (p=0.056).

**Conclusion:** Important to transmission and disease course, oral viral replication and persistence showed clear relationships with select symptoms, early Ig responses, and gender during early infection. N-antigen cross-reactivity may reflect mimicry of structurally analogous host proteins.

## Introduction

SARS-CoV-2 emergence became the source of the largest coronavirus-driven pandemic. This positive-strand RNA virus, and COVID-19 etiologic agent, infects and replicates in the upper respiratory tract, respiratory mucosa, oral mucosa and salivary glands^1^. The presence of Ace2 receptor+ oral cells, SARS-CoV-2 RNA detection, and detection of virus capable of inducing cytoplasmic effect suggests that oral viral replication and production occur^1^. SARS-CoV-2 was consistently detected in unstimulated whole mouth fluid (WMF), oropharyngeal WMF, and gingival crevicular fluid ^2–6^. The CDC and the European Center for Disease Prevention and Control recommend nasal/oral swabs or saliva testing^7^. Nasopharyngeal swab reverse transcriptase-polymerase chain reaction (NP-RT-qPCR) is the gold standard for SARS-CoV-2 detection, however, adequacy of harvested material and collection time relative to disease onset can result in low methodologic sensitivity^8, 9^. Meta-analysis demonstrated high concordance, 92.5% (95%CI: 89.5-94.7), between saliva and nasopharyngeal/oropharyngeal swabs (NPS/OPS) with lower sensitivities in saliva than nasopharyngeal/oropharyngeal swabs, 86.5% (95%CI: 83.4-89.1) and 92.0% (95%CI: 89.1-94.2), respectively^7^.

Cochrane assessment of SARS-CoV2 antigen tests (n=48), including lateral flow assays (LFA), demonstrated varied sensitivity between symptomatic and asymptomatic participants with highest sensitivity closest to symptom onset^10^. While LFA-assessed oral SARS-CoV-2-targeted immune responses can reflect systemic responses^11^, oral biomarkers as prognostic COVID-19 indicators have not been significantly explored. Here, RT-qPCR, LFA, and immunoblot were used for oral SARS-CoV-2 detection. Longitudinal assessment of symptomatic participants suggested oral viral persistence. Disease severity and symptoms were associated with oral SARS-CoV-2 host responses and viral presence. Collectively, these findings provide novel insights to oral markers of prognosis, persistence, and transmission.

## Materials and Methods

### Enrollment

47 ambulatory participants provided IRB-approved written informed consent. Participants were part of the 1) longitudinal symptomatic group (n=17), at entry SARS-CoV-2 NP-RT-qPCR+ with stratified symptoms (mild, moderate, severe) based on NIH COVID-19 treatment guidelines, 2) blinded asymptomatic seropositive/seronegative cross-sectional group (n=30) ^12^, 3) Archived, preCOVID-19 era saliva and throat wash (TW)^13^.

### Biospecimens

NPS in transport medium, saliva, and TW were collected at the same visit. Unstimulated WMF (saliva) and TW gargle with 10 ml of saline were collected. Blood was collected by venipuncture for serology. Samples were transported, stored at 4°C, and processed within 24 hours.

### Nucleic acid extraction and RT-qPCR

Virus was heat/chemically inactivated. RNA was extracted using Trizol (Life Technologies) and reverse transcribed using Superscript III (Life Technologies), according to manufacturer’s instructions. First strand synthesis was primed with random hexamers (Life Technologies) and reactions used in SYBRGreen-based qPCR. The SARS-CoV-2 genome was targeted using 3 regional primer sets described in Figure 3A^14, 15^. A SARS-CoV-2 reference RNA-based (ATCC VR-3276SD) standard curve allowed absolute copy number quantitation. Subgenomic RNA (sgRNA) detection across viral junctions was performed using the forward primer, CoV25UTR with gene-specific reverse primers (**Figure 3A**). Percent sgRNA was calculated using the equation: 2^-(sgRNA Ct – NRNA Ct)/100. Positive reactions were treated with ExoSapIt according to manufacturer’s instructions (USB, Cleveland, Ohio) and Sanger sequencing performed using SARS-CoV-2-specific primers (Eton Bioscience, Research Triangle Park, NC).

### Recombinant His-tagged SARS-CoV-2 Nucleocapsid (N-antigen)

E. coli BL21 (DE3) bacteria were plasmid transformed with SARS-CoV-2 N coding region ligated in frame to the pET30 6X histidine-tagged coding region. After IPTG (1 mM) induction, bacterial pellets were lysed and protein purified with NTA-Ni agarose beads (Life Technologies). Eluates were subjected to SDS-PAGE followed by Coomassie staining for retrieval and protein purity.

### Ectopic expression of SARS-CoV-2 proteins in oral keratinocytes

Full-length subgenomic PCR products encoding SARS-CoV-2 S, E, M or N were generated from SARS-CoV-2+ salivary total RNA. cDNAs were cloned into pCMV-myc and transfected into immortalized human oral keratinocytes (NOK) using Fugene-6 (Promega). Forty-eight hours post transfection, media was subjected to SDS-PAGE/ immunoblot to detect myc-tagged proteins.

### Immunoblot

Total protein was isolated using Trizol according to the manufacturer’s instructions. Protein pellets were resuspended in sample buffer and proteins electroblotted to PVDF membrane. Blots were blocked followed by primary antibody incubation, incubation with secondary HRP-conjugated antibody, then protein detection by ECL Prime (GE) and imaging using GE ImageQuant LAS4000.

### Lateral Flow Assay

SARS-CoV-2 (N-antigen, anti-SARS-CoV-2 spike protein IgG/IgM) were detected in TW/saliva using LFA cartridges according to manufacturer’s instructions (BioMedomics, Research Triangle Park, NC)^16, 17^. Bands were visualized and quantitated using ImageJ. Control bands for each detection strip were used to normalize antigen-specific bands and IgG/IgM intensity. Quantitation was determined relative to preCOVID-19, archived saliva/TW signal.

### NCBI Structural analysis

The SARS-CoV-2 N-antigen N-terminal RNA binding domain (RBD) crystal structure (PDB ID: 6M3M) was compared to other publicly available crystal structures in the Molecular Modeling Database (MMDB) using Vast+ (https://www.ncbi.nlm.nih.gov). 3D structures of superimposed biological assemblies were visualized using the web-based 3D viewer, iCn3D version 4.3.1^18^.

### Statistical methods

Given small sample sizes, non-parametric tests were used to test correlations and compare data. The Kendall rank correlation test assessed the existence of monotonic relationships of ordinal or continuous variables using the ranks of the data. The Mann–Whitney U test was used where the independent variables were binary and determined whether the two groups were from the same population when the groups were independent. Mann–Whitney U and Wilcoxon signed rank tests were used to compare response variables between two groups. When groups were dependent, Wilcoxon signed rank tests assessed differences in the medians of matched groups.

## Results

### Assessment of the SARS-CoV-2 N-Antigen Lateral Flow Assay

N-antigen LFA was validated using lysates from immortalized normal oral keratinocytes (NOK) transfected with a myc-tagged SARS-CoV-2 N-antigen expression vector. N-antigen was detected by anti-myc antibody immunoblot (**Figure 1A**) or LFA (**Figure 1B**). Bacteria-produced, his-tagged N-antigen was purified (**Figure 1C**), used for N-LFA quantification, and migrated at 55 KD, matching the predicted molecular weight of SARS-CoV-2 N-antigen, while pET30 vector negative control ran at ∼9 KD (**Figure 1C**). LFA limit of detection was determined using two-fold serial dilutions of preCOVID-19 saliva spiked with recombinant N-antigen and detected his-tagged N-antigen at 43-680 picograms (pg). A faint band at 85 pg and robust detection >170 pg demonstrates LFA’s semi-quantitative nature (**Figure 1D**). No band was detected in saliva spiked with pET30 recombinant protein, indicating signal specificity. LFA allowed longitudinal detection of N-antigen in saliva from a representative NP-RT-qPCR+ participant at symptom onset 14, and 28 days, suggesting active persistent infection, while signal was undetected in two PreCOVID-19 saliva samples (**Figure 1E).**

**Figure 1.**
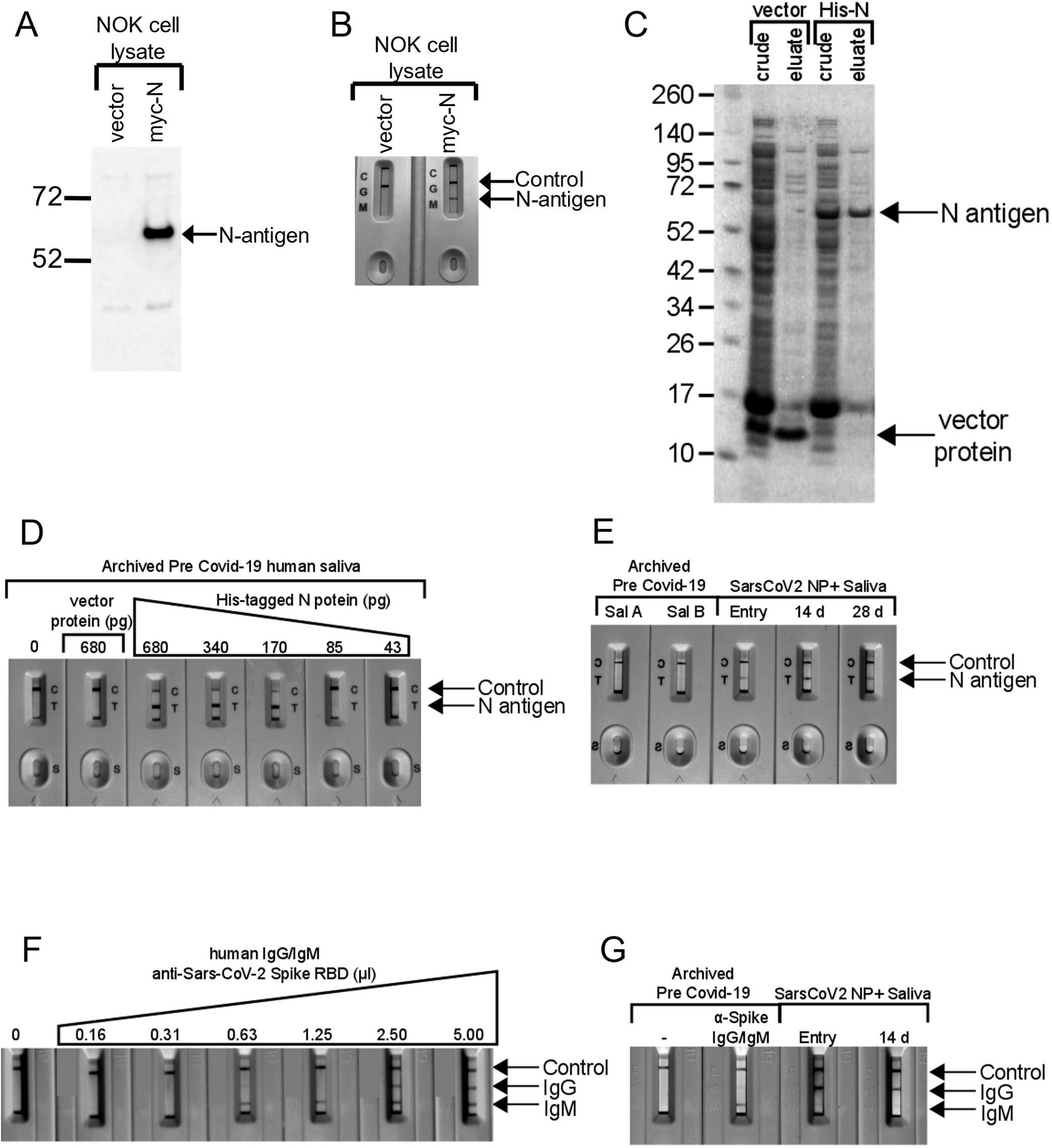
Assessment of Lateral flow assays for detection of SARS-CoV-2 N-antigen and anti-SARS-CoV-2 spike RBD domain targeted antibodies. **A.** Expression of myc-tagged N-antigen in NOK cells by immunoblot. Vector lane indictates cells transfected with the empty plasmid, pCMV-myc. N-antigen detected with anti-myc antibody is indicated by the arrow. **B.** LFA detected N-antigen in the lysates of NOKs transfected with myc-tagged N expression plasmid. Both, control and N-antigen bands, are indicated. Only the control band is detected in lysates from cells transfected with empty vector. **C.** Expression and purification of His-tagged SARS-CoV-2 N-antigen in E. coli. A band consistent with the size of N-antigen (55 kD) can be seen in crude lysates from cells transformed with His-tagged, N-antigen expression plasmid. This band is eluted from Ni-beads during purification. His-tagged protein expressed in bacteria transformed with empty vector (pET30) served as a negative control. **D.** Dilution series of His-tagged N-antigen in preCOVID-19 saliva and detection using Biomedomics N-antigen LFA. **E.** Detection of N-antigen in saliva of preCOVID-19 and symptomatic SARS-CoV-2 NP+ subjects at entry and post entry 14D and 28D by LFA. The detection of N-antigen band is indicated. **F.** Two-fold dilution series of anti-SARS-CoV-2 Spike RBD (S-RBD) domain IgG/IgM antibodies in pre COVID-19 saliva and detection using Biomedomics IgG/IgM lateral flow assay cartridges. Control, anti-S-RBD IgM and S-RBD IgG are indicated. The 0 lane indicates unspiked, preCOVID-19 saliva only.

### Assessment of the SARS-CoV-2 Anti-Spike RBD IgM/IgG Lateral Flow Assay)

Recombinant anti-Spike RBD IgG/IgM in LFA assays detected anti-SARS-CoV-2 Spike-specific IgM and IgG oral immune responses. Two-fold dilution allowed immunoglobulin LFA validation. Mixtures of Spike RBD-specific recombinant human IgM/IgG in preCOVID-19 saliva detected analytical sensitivity down to 0.63 microliters for IgM, however, IgG was undetected after a single 2-fold dilution (**Figure 1F**). Archived saliva, spiked with anti-SARS-CoV-2 antibody demonstrated IgM signal but not IgG, perhaps reflecting IgM detection of a pre-2019 human coronavirus. Unspiked demonstrated no signal (**Figure 1G**). A symptomatic participant demonstrated IgG at baseline and at 14 days but not IgM (**Figure 1G**).

### Symptomatic and Asymptomatic Assessment

Ambulatory groups who participated in survey and biospecimen collection were: 1) deidentified-symptomatic, NP-RT-qPCR+, 2) blinded-asymptomatic seropositive or 3) blinded-asymptomatic seronegative (**Figure 2A**). Symptomatic participants (n=17) were assessed at baseline, 14 and 28 days. The mean age was 40.2 years with equivalent gender distribution. The cohort was 64.7% Caucasian, 20% Hispanic/Latino, and 6.4% each of Black and Asian participants (**Figure 2B**). RT-qPCR targeted three SARS-CoV-2 genomic regions (orf1, E and N) in saliva/TW. Known amounts of N viral RNA were detected by standard curve, allowing absolute copy number quantification, that was more sensitive than relative quantitation of E/orf1(**Figure 3F**). All full-length and subgenomic (sgRNA) viral RNAs are amplified by 2019-nCoV_N1-F/2019-nCoV_N1-R allowing relative quantitation (**Figure 3A**). Reactions containing >10 copies were consistently reproducible (**Figure 3B, F**). PreCOVID-19 saliva/TW RT-qPCR, did not produce signal >10 copies/reaction. Oral samples containing >10 copies/reaction were considered SARS-CoV-2 RNA positive. Using these criteria, 59% of saliva samples (10/17) and 64% (9/14) of TW samples from the symptomatic group were SARS-CoV-2 RNA+ (**Figure 3F**).

**Figure 2.**
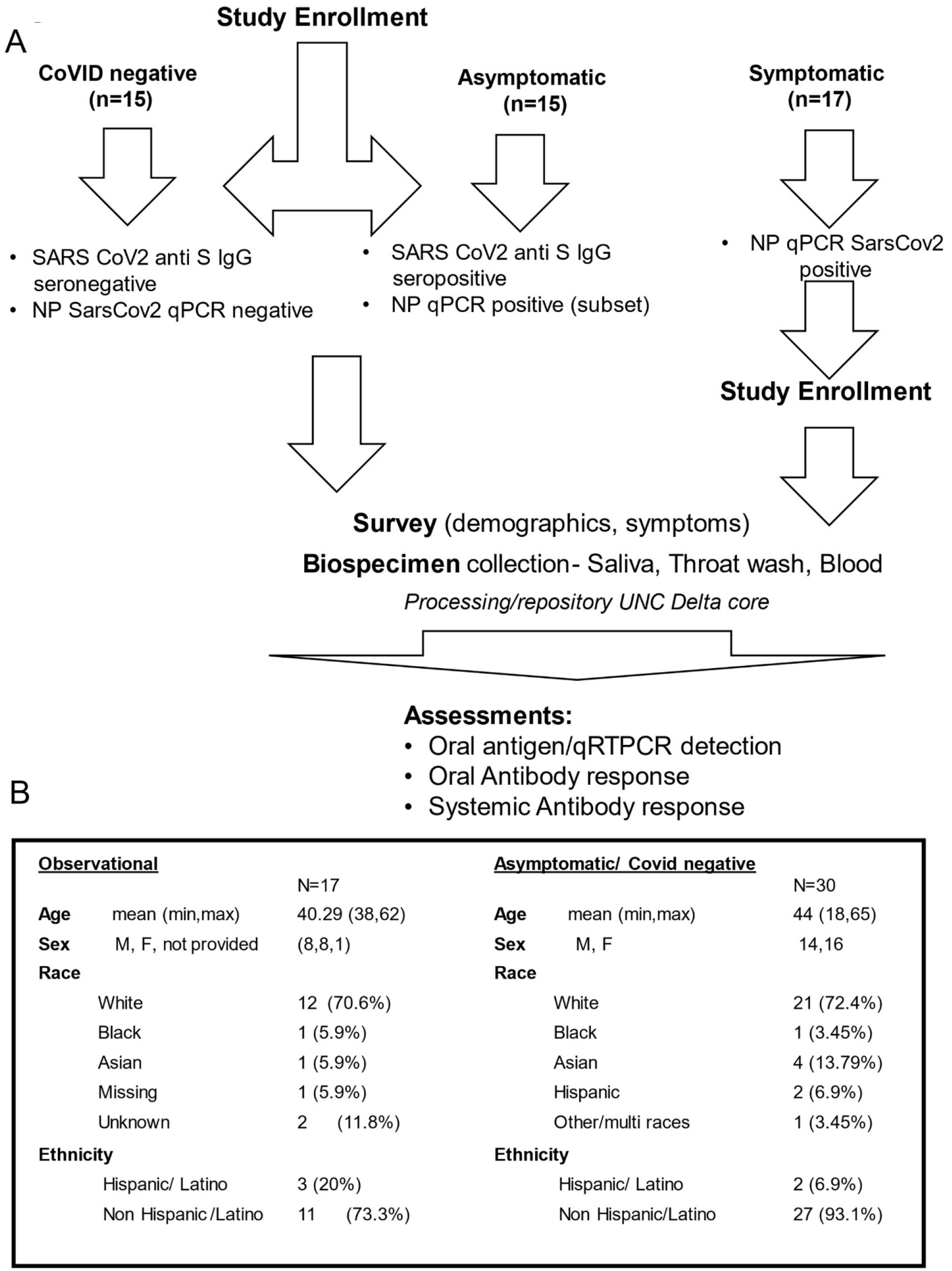
Study Schema and Asymptomatic/symptomatic Participant Demographics. **A.** Enrollment inclusion criteria for subjects who were COVID-19 negative (n=15), had asymptomatic COVID-19 (n=15), or had symptomatic COVID-19 infection (n=17) are listed within the figure. Subsequent to enrollment, participants answered surveys and provided biospecimens (saliva, TW, blood) that were assessed for SARS-CoV-2 detection and antibody responses. B. Symptomatic SARS-CoV-2 NP+ and asymptomatic participant demographics are indicated.

**Figure 3.**
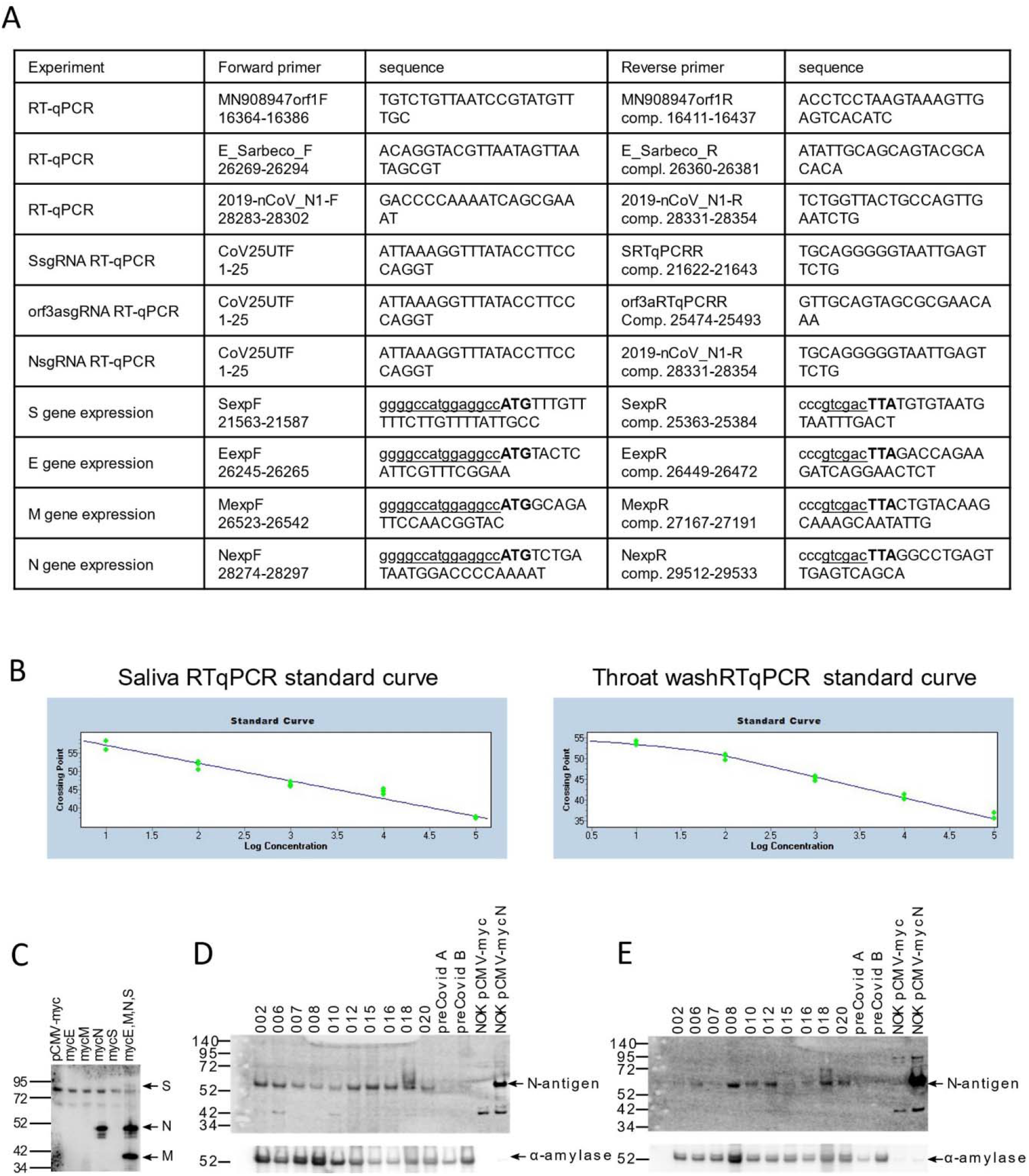
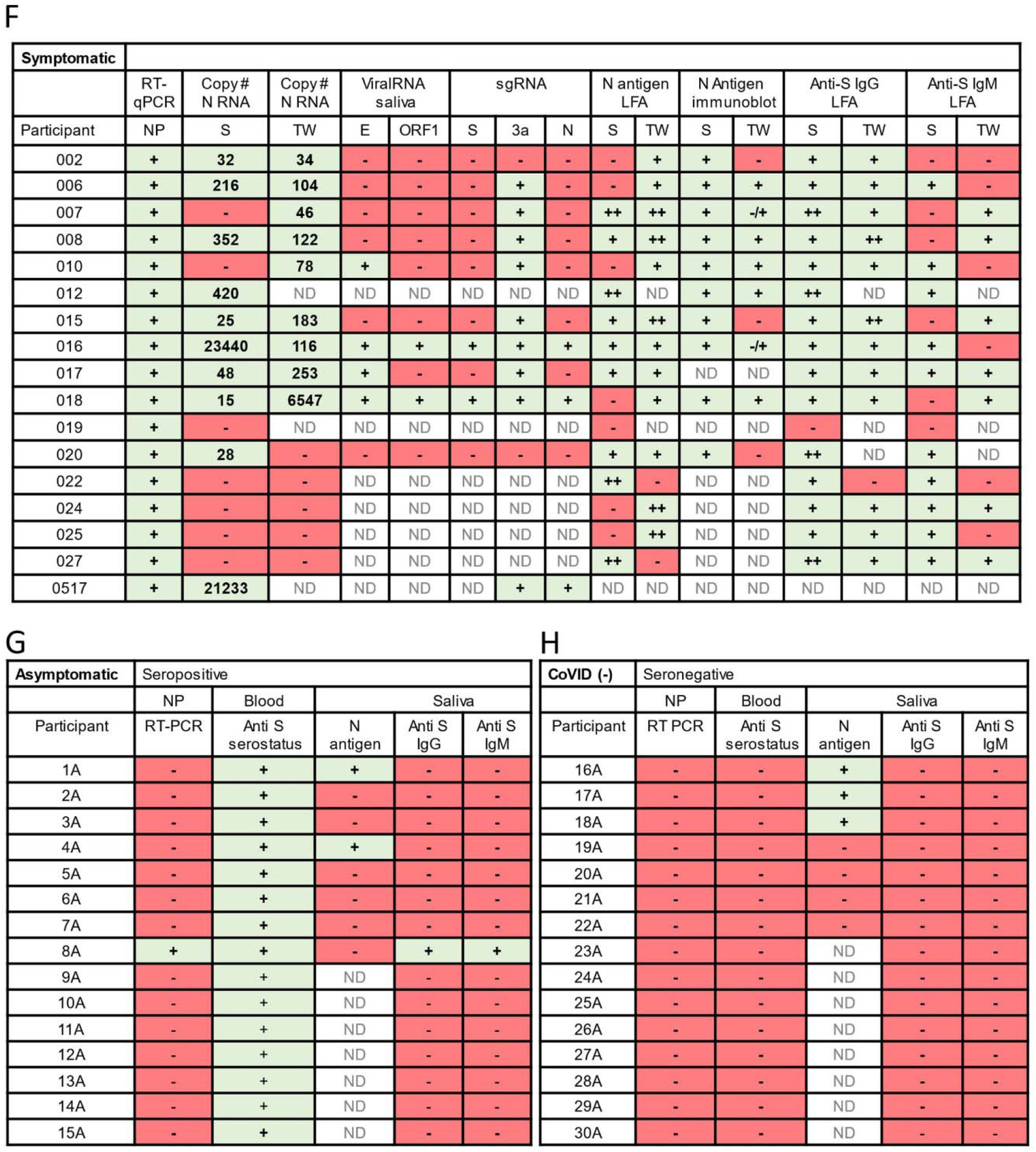
Oral virologic characterization of study participants at entry. All subjects were assayed by qPCR for detection of viral RNA in the nasopharynx. Saliva/TW was assayed by qPCR for viral RNA and for LFA-based detection of N-antigen, anti–spike IgG and anti-spike IgM. **A.** Oligonucleotide sequences for primers used for RT-qPCR and production of cDNA for construction of expression plasmids. E_Sarbeco_F and E_Sarbeco_R have been described. The sequences of 2019-nCoV_N1-F and 2019-nCoV_N1-R, were obtained from the CDC. The SARS-CoV-2 reference sequence GenBank:MN908947 was used to determine oligonucleotide sequences and map coordinates. SfiI (ggccatggaggcc) and SalI (gtcgac) restriction enzyme sites used for cloning into the expression vector, pCMV-myc, are underlined in expression primers. The start codon and stop codons in expression primers are in bold. **B.** Standard curves were generated by RT-qPCR using know copy numbers of SARS-CoV-2 RNA and primers targeting the nucleocapsid (N) coding region to quantitate viral RNA isolated from patient saliva and TW. **C.** Immunoblot was used to detect SARS-CoV-2 structural proteins in cell culture media obtained from NOKs transfected with individual expression vectors encoding myc-tagged, SARS-CoV-2 structural proteins (S, E, M and N) or cotranfected with all myc-tagged expression vectors. Medium from cells transfected with empty vector (pCMV-myc) was used as a negative control. Proteins were detected using myc-specific antibody (mouse anti-myc, sc-40, Santa Cruz Biotechnology). **D and E.** Protein from subset of saliva and TW samples (respectively) was used for the detection of viral proteins. Immunoblot detection of SARS-CoV-2 N-antigen was assessed in a subset of symptomatic individuals (n=10) at baseline (top panel) using N-specific antibody (PA5-114448, LifeTechnologies, Inc). Detection of salivary alpha-amylase (anti-amylase, A8273, Sigma-Aldrich) served as a loading control (bottom panel). Saliva or TW from archived preCOVID-19 patients A and B were included as negative controls. Cell culture medium from NOK cells transfected with myc-tagged SARS-CoV-2 N-antigen expression construct was used as a positive control. Cell culture medium from NOKs transfected with empty vector was included as a negative control. N-antigen was detected at 55 KD and alpha amylase, the loading control, was detected at 58.4 KD. **F.** Table of symptomatic subjects (n=17) who were all nasopharynx RT-qPCR positive. RT-qPCR and LFA was also performed on saliva and TW of these subjects. Viral RNA copy number (molecules of virus RNA) in 25 microliters of saliva or TW was determined using N-specific primers. Green highlight indicates a positive result. Red highlight indicates a negative result. ND indicates not determined. **G.** Asymptomatic participants (n=15) were all anti spike IgG positive in their blood. **H.** COVID-19 negative participants (n=15) were anti-spike negative and nasopharynx negative.

Oral N-antigen was assessed by immunoblot. Media from NOKs, transfected with myc-tagged SARS-CoV-2 structural protein expression vectors, was subject to myc-specific immunoblot, demonstrating readily detectible N-antigen. Detection of myc-tagged spike (S) and matrix (M) proteins in the medium depended on N expression suggesting virus-like particle (VLP) production (**Figure 3C**). Immunoblot analysis, using a commercially available SARS-CoV-2 N-antigen antibody, detected salivary N-antigen at the expected migration of 55 KD in 10/10 subjects assayed, similar to myc-tagged recombinant N-antigen. pCMV-myc transfected cell medium and preCOVID-19 saliva samples, A and B, were negative controls (**Figure 3D**). TW immunoblot detected N-antigen in 7/10 subjects with 2/7 displaying faint bands (**Figure 3E**). The salivary protein, alpha-amylase served as a loading control. There was striking concordance between NP positivity (reference), salivary N-antigen immunoblot detection, and IgG detection - salivary (93%)/TW (100%) (**Figure3F**). IgM concordance with NP-RT-qPCR+ status was 75% in saliva and 50% in TW.

The asymptomatic group (n=30) was composed of seropositive (n=15) and seronegative/uninfected (n=15) participants. The mean age was 44, with 14 males and 16 females and was 72.4% Caucasian, 4% Asian, 7% Hispanic and 3.4% Black. Of 15 seropositive participants, only one, subject 8A, was NP-RT-qPCR+, IgG positive, and IgM positive in the saliva (**Figure 3G**). Two others (1A and 4A) within the seropositive-asymptomatic group were LFA positive (**Figure 3G**). Among seronegative subjects, salivary IgG and IgM were not detected however, 3/7 subjects demonstrated N-antigen by LFA (**Figure 3H**). To determine potential targets capable of cross-reactive binding, host salivary proteins structurally analogous to N were assessed (**Figure 4**). The vector alignment search tool, VAST+, localized macromolecular structures similar in 3D shape to SARS-CoV-2 N-antigen^19^. VAST+ detected N-antigen complete or partial matches, that included SARS-CoV-1 and human RNA binding proteins Line1Orf1p, hnRNP H, RBM7, and 17S U2 snRNP. Salivary proteome members RBM7 and 17S U2 snRNP, demonstrated 29aa structural alignment at 1.47 angstroms and 27aa structural alignment at 1.52 angstroms, respectively. Line1ORF1p (19aa at 1.00 angstroms) and hnRNP H (28aa at 1.50 angstroms) have salivary proteome isoforms^20^. SARS-CoV-1 served as a reference control demonstrating structural alignment of 95aa at 0.58 angstroms (**Figure 4**).

**Figure 4.**
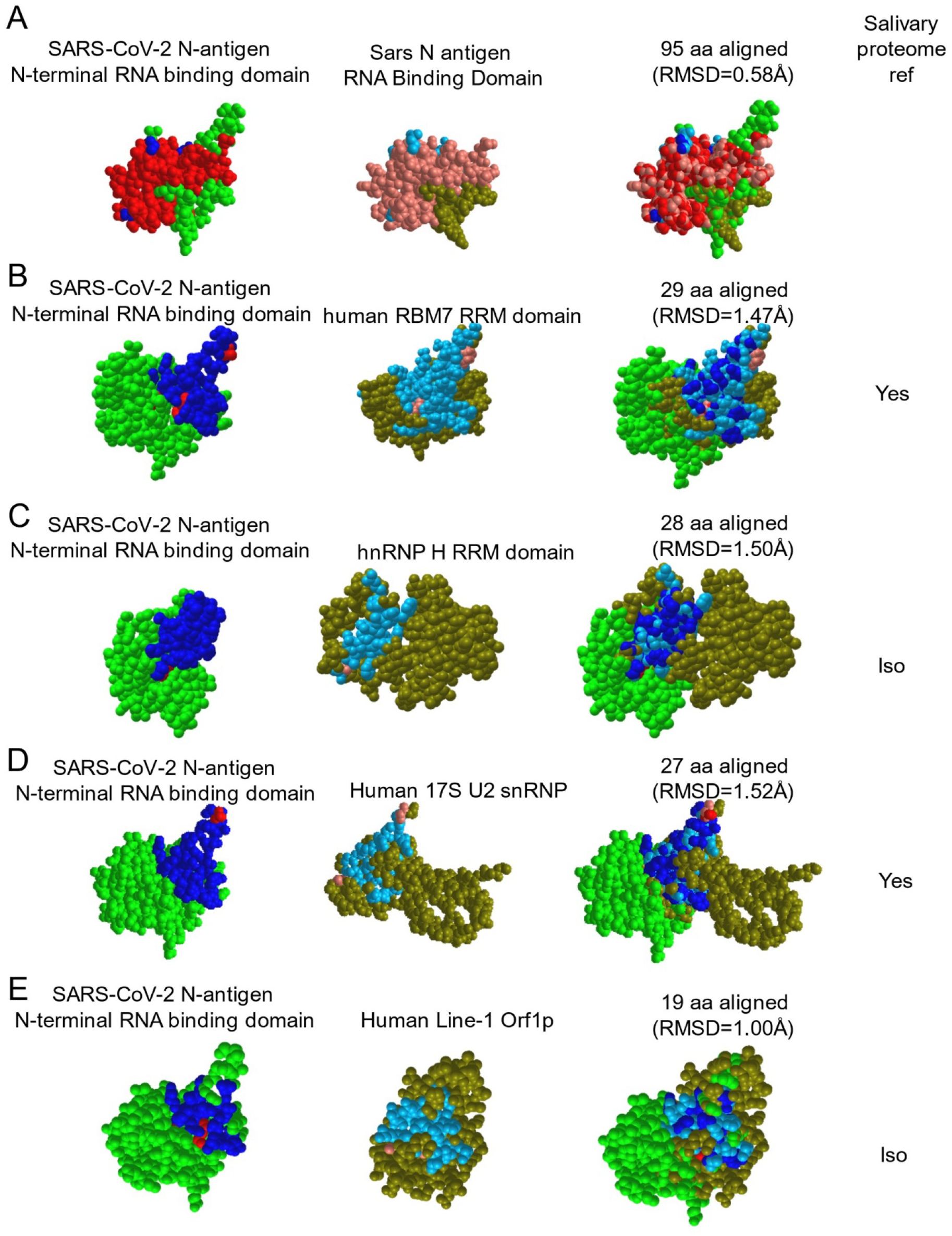
The SARS-CoV-2 nucleocapsid mimics host RNA binding proteins that are expressed within the salivary proteome and may be responsible for cross reactivity in LFA assays. VAST+ was used to generate RNA binding protein structures demonstrating 3D similarity to the SARS-CoV-2 N-antigen. Simultaneous alignment generated molecular protein pairs between N-antigen (first column) and host RNA binding proteins (second column). Non-aligned N-antigen amino acids are rendered in green, conserved aligned amino acids are rendered in red and aligned, non-conserved amino acids are rendered in blue. Non-aligned host protein amino acids proteins are rendered in olive green, conserved aligned amino acids are rendered in pink and aligned, non-conserved amino acids are rendered in light blue. The spatial arrangement of overlapping amino acids can be seen in the merged sequence alignment (third column). The number of structurally aligned amino acids (conserved and non-conserved) is shown. The root mean square deviation (RMSD) is used as a measurement between atoms in the backbone of the molecular structures and is listed in angstroms above each merged image and detection of the host protein within the salivary proteome is displayed as yes in the fourth column while detection of a related isoform within the salivary proteome is displayed as ISO.

Detection of sgRNAs, arguably only produced during active viral replication, confirmed TW LFA positivity (**Figure 5,3F**). Detection of overlapping junction sequences between the end of the 5’ UTR and the beginning of the S, orf3a, or N coding regions in oral fluids (**schematic, Figure 5A**) was confirmed by direct sequencing of qPCR products shown as qPCR+/number assayed - 20% (2 /10), 82% (9/11), and 27% (3/11) subjects, respectively (**Figures 5B-D**). No RT-qPCR signal for sgRNAs were detected in preCOVID-19 RNA samples/water negative control. Overall, 90-100% of those who were RNA+ in oral fluids (sal/tw) demonstrate orf3a sgRNA, were LFA N-antigen positive, immunoblot confirmed, and IgG positive (**Figure 3F**).

**Figure 5.**
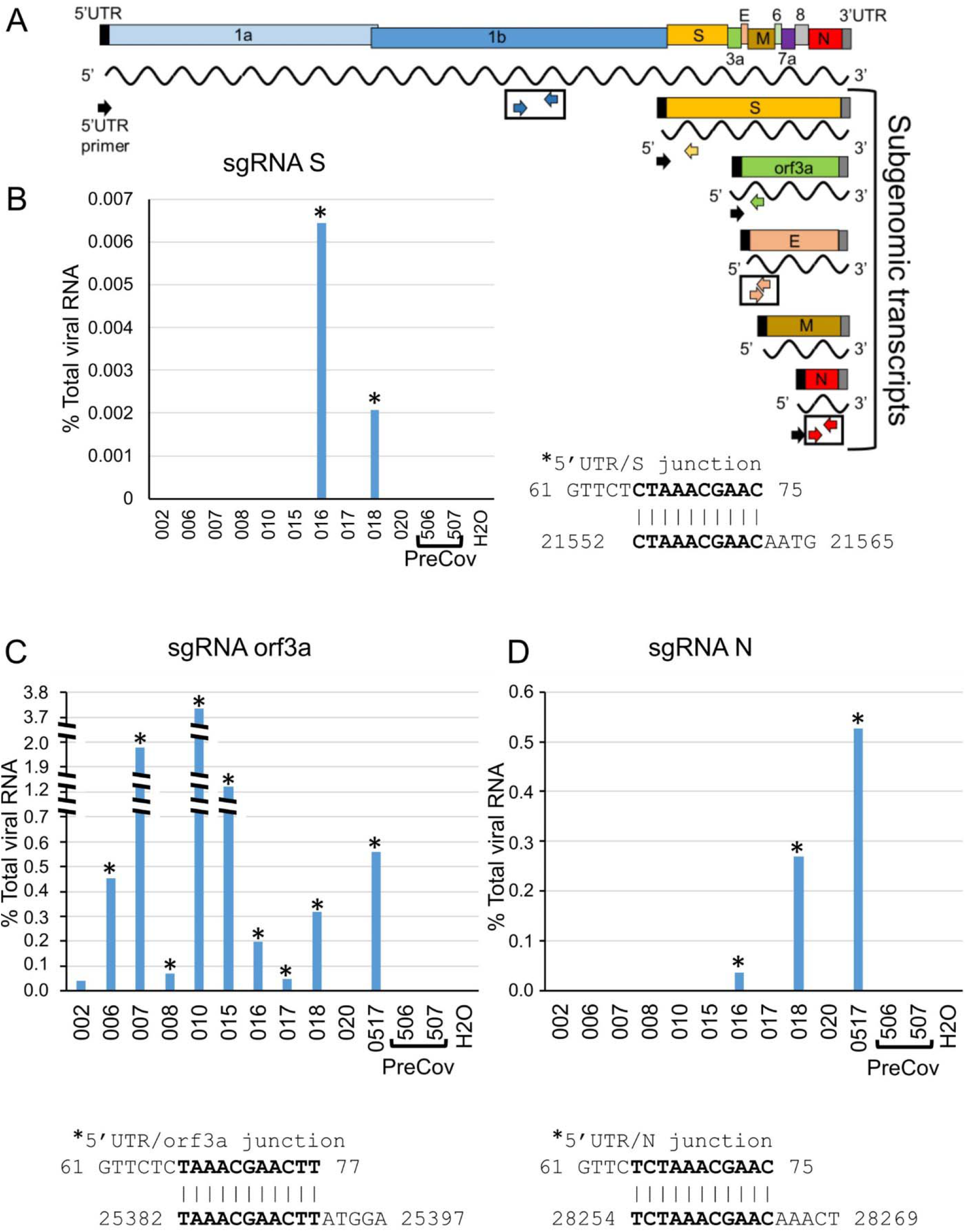
SARS-CoV-2 subgenomic RNAs (sgRNA) are detected the saliva of symptomatic COCID patients. **A.** Schematic diagram showing the RNA genome of SARS-CoV-2 (∼30 kb) and the subgenomic RNAs encoding major viral structural proteins (S, E, M and N) as well as orf3a. Detection of SARS-CoV-2 RNA by RT-qPCR was performed using primer pairs corresponding to orf1 (blue arrows), E (peach arrows) and N (red arrows) coding regions. SgRNA was detected using a forward primer corresponding to the 5’UTR (black arrow) and either S-specific (yellow), orf3a-specific (green) or N-specific (red arrow) reverse primers. The RT-qPCR signal generated with N1 primers (red arrows) were used to calibrate levels of sgRNA. **B, C and D.** The percent levels of S, orf3a or N sgRNA determined using the 5’UTR primer and the specific sgRNA reverse primer are shown. Percent sgRNA levels relative to total viral RNA levels are determined as follows: 2^-(sgRNA Ct/N Ct) X 100. Direct sequencing of RT-qPCR amplified products was performed to determine the junction sequence of the 5’UTR and the coding region of each sgRNA. Sequencing information obtained from samples is indicated with an asterisk. The overlap sequence between the 5’UTR and each specific sgRNA is indicated.

### Viral metrics in the oral fluids over time and between genders

LFA test and control band intensity was measured by ImageJ to provide relative measures of N-antigen and immunoglobulin presence in oral fluids. An insignificant downward trend in salivary N-antigen detection from baseline to 28 days, suggested persistence in oral fluids (Wilcoxon Signed Rank Test) (**Figure 6A**). While quantitatively less N-antigen was detected in symptomatic TW at 28 days compared to baseline (relative band intensity, median=2070 vs 2310), this decrease was not significant (Wilcoxon Signed Rank Test, p=0.9). In the symptomatic group, significant differences were detected between preCOVID-19 archived saliva, baseline, and 28 days (Wilcoxon Rank Sum Test, p=0.003) (**Figure 6B**). At baseline, 56% of the symptomatic were N-antigen positive in saliva and 91% were N-antigen positive in TW by LFA. Using NP-RT-qPCR as the gold standard, LFA assay sensitivity was 0.563 (CI, 0.299, 0.802) in saliva and 0.91 (CI, 0.587,0.998) in TW (**Figure 6C**). Salivary IgG levels were significantly different between baseline and preCOVID-19 archived samples (Wilcoxon Rank Sum Test, p=0.005). IgG trended upward over time and IgM levels, approximately a log lower than the IgG levels, remained the same over 28 days (p=0.625) (**Figure 6A**). While gender differences were not detected in oral N-antigen or TW IgG, females demonstrated higher levels of IgM than males (Mann-Whitney U test, p=0.056) (**Figure 6D**).

**Figure 6.**
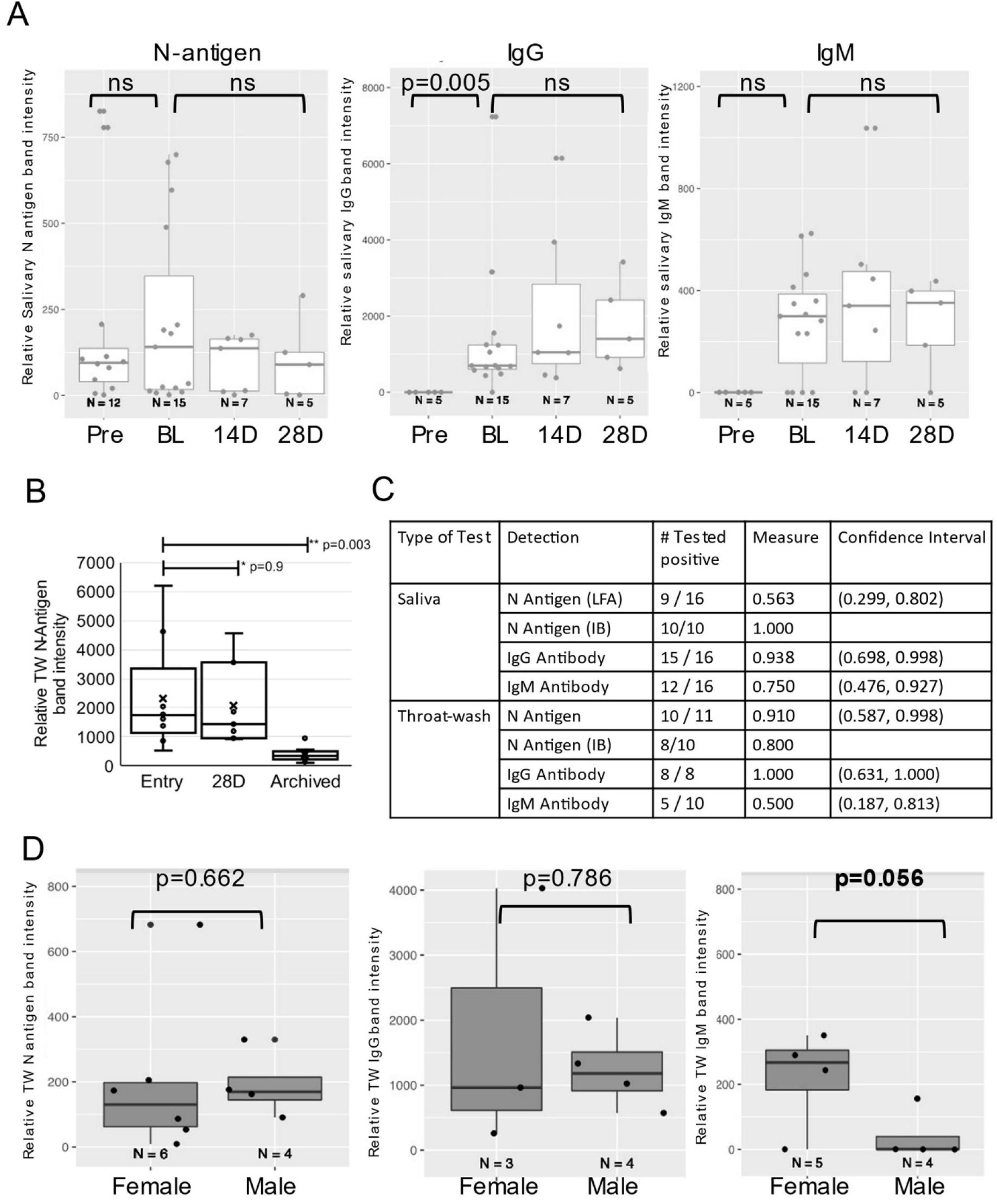
Longitudinal semi-quantitative analysis of SARS-CoV-2 N-antigen and anti-SARS-CoV-2 specific IgG/IgM antibodies in oral fluids of infected individuals demonstrates persistence and gender differences. **A.** Detection of N-antigen (left panel), anti-Spike IgG (middle panel), and anti-Spike IgM (right panel) in TW of symptomatic subjects at entry (BL), 14 days (14D), and 28 days (28D) post entry compared to preCOVID-19, archived (Pre) TW. **B.** LFA detection of N-antigen in TW of symptomatic subjects at entry (BL) and 28 days (28D) post entry compared to preCOVID-19, archived TW. The Wilcoxon Rank Sum test assessed difference between preCOVID-19 and entry and Wilcoxon Signed Rant Test assessed significant differences between matched entry and 28 day samples. **C.** Saliva and TW N-antigen, anti-Spike IgG and anti-Spike IgM LFA assay sensitivity was determined relative to NP-RT-qPCR+ results in symptomatic participants. **D.** Detection of N-antigen (left panel), anti-Spike IgG (middle panel), and anti-Spike IgM (right panel) in TW of female vs male symptomatic subjects at entry demonstrates differences in relative IgM levels.

### SARS-CoV-2 oral outcomes, COVID-19 symptoms and symptom severity, and oral persistence

Self-reported COVID-19 symptom severity at baseline (absent, mild, moderate, or severe) and presence of SARS-CoV-2 oral N-antigen or antibody in saliva/TW were assessed. COVID-19 symptom (weakness, muscle ache, nausea, loss of taste/smell and upper respiratory tract symptoms which encompassed cough, shortness of breath, sore throat, nasal obstruction, nasal discharge) presence or absence at baseline was reported by participants as yes/no. Kendall Rank Correlation tests determined associations between severity of cough and fatigue with salivary IgM (p=0.008 and 0.016 respectively) (**Figure 7A and B**). At baseline, salivary IgM was consistently associated with weakness (p=0.037), muscle ache (p=0.019), nausea (p=0.005) and upper respiratory symptoms (p=0.017), but not loss of taste/smell (p=0.458) (Mann-Whitney U test). Oral N-antigen and IgG detection were not associated with COVID-19 symptoms in oral fluids (Mann-Whitney U test) (**Figure 8A)**. While longitudinal assessment of oral antigen and immunoglobin responses detected no clear directional trends, there was consistent detection of IgM, IgG, and oral N-antigen at baseline and over time (n=11) (**Figure 8B**).

**Figure 7.**
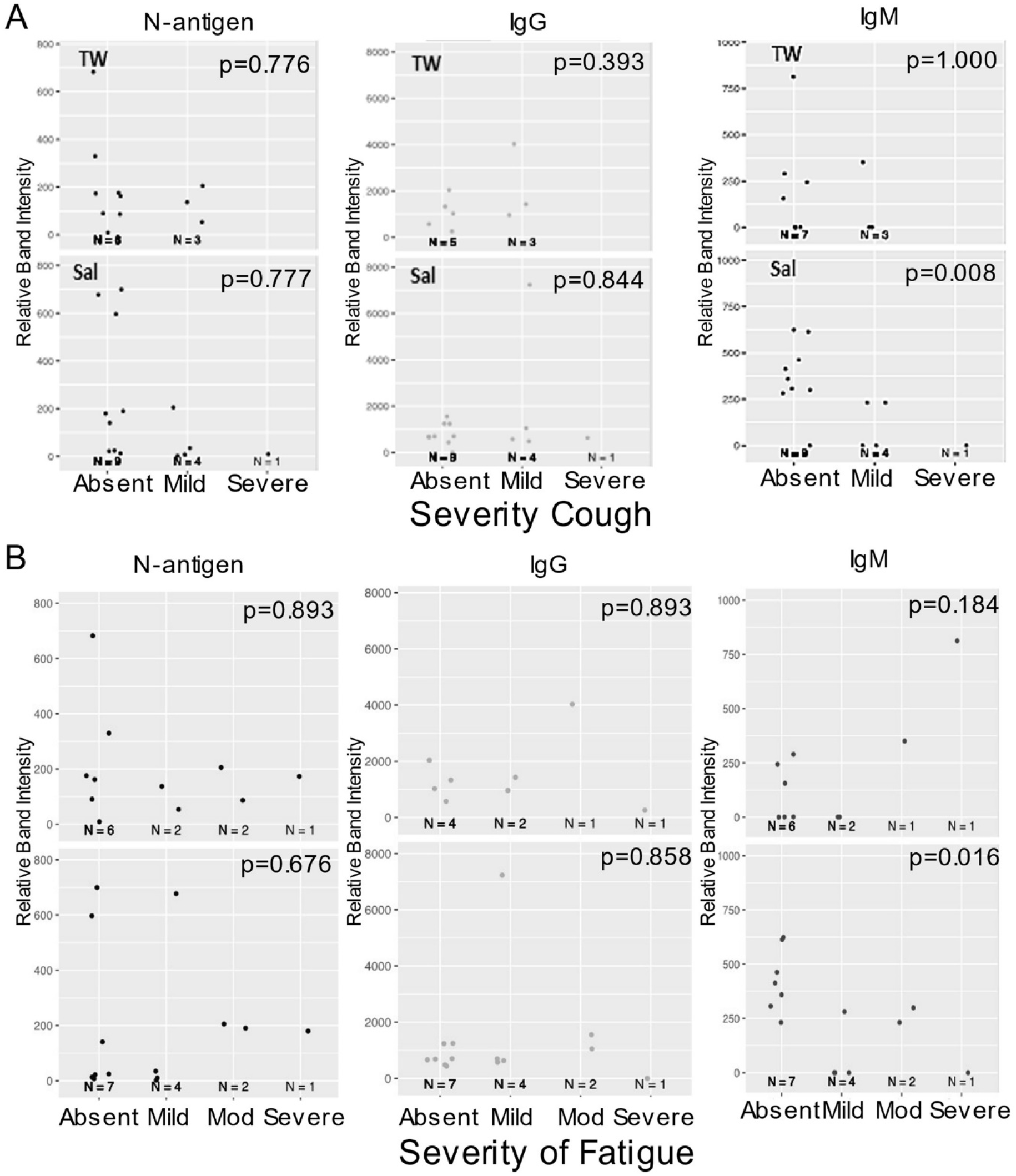
Salivary IgM predicted a more favorable disease course. The relationship between symptom severity and semi-quantitative SARS-CoV-2 protein and anti-spike antibody levels in saliva and TW of symptomatic infected individuals with cough and fatigue was assessed at baseline using LFA. Kendall Rank Correlation test detected a decreasing relationship between salivary IgM detection and the severity of fatigue and cough.

**Figure 8.**
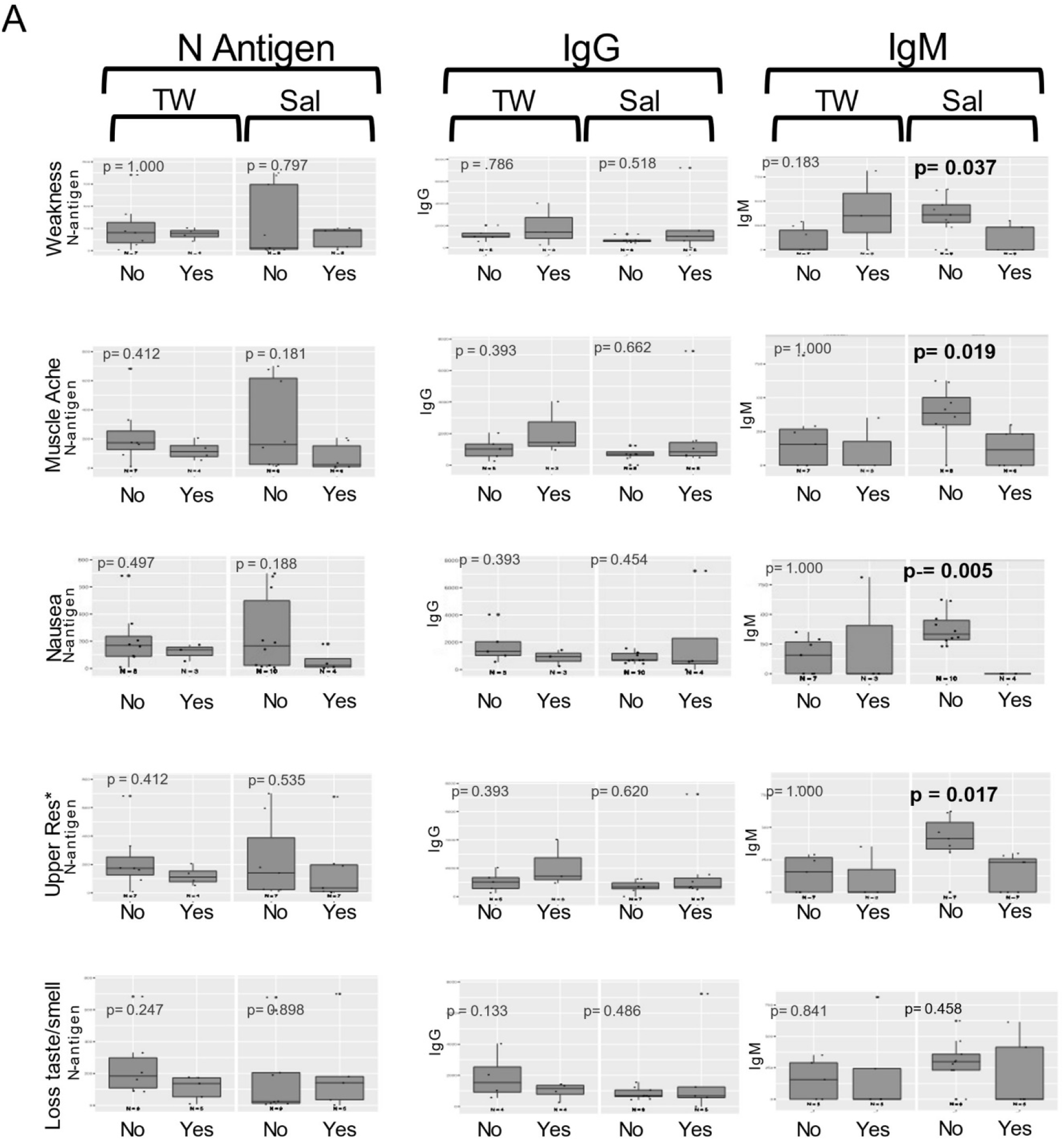
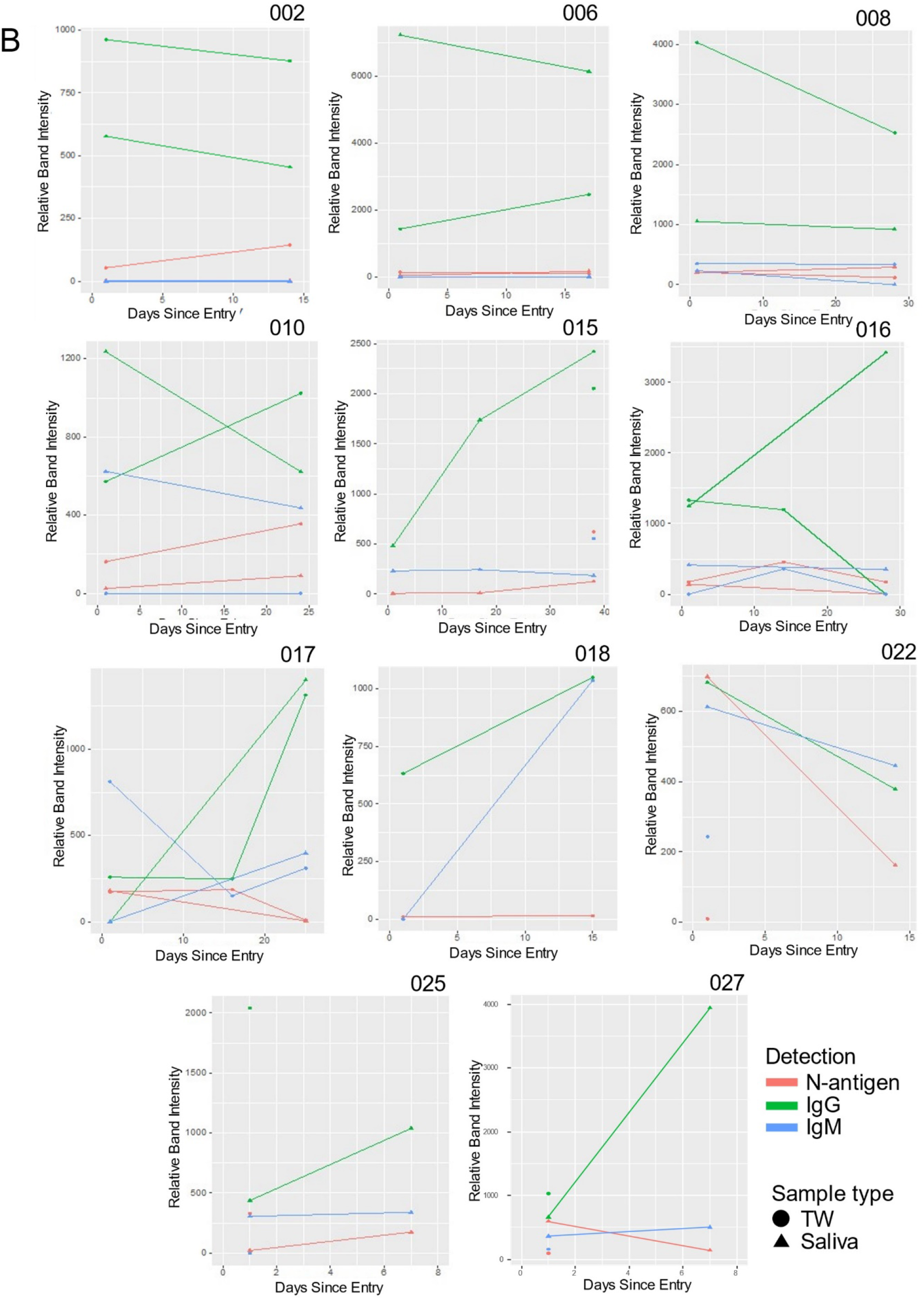
Salivary IgM was associated with the absence of most symptoms. **A**. The relationship between symptom presence (weakness, muscle ache, nausea, loss of taste and smell and upper respiratory symptoms [including cough, shortness of breath, sore throat, nasal obstruction (stuffy nose), nasal discharge [runny nose]) and semi-quantitative SARS-CoV-2 N-antigen and anti-spike antibody levels in saliva and TW of symptomatic infected individuals was assessed at baseline as determined by LFA. The Mann-Whitney U test detected a relationship between the absence of symptoms and higher oral IgM levels in saliva. **B**. Longitudinal oral virologic characterization of symptomatic participants detects viral persistence with concurrent antibody responses suggesting presentation post-acute infection. Saliva (triangle) and TW (circle) were assayed by LFA for detection of N-antigen (red), anti –spike IgG (green) and anti-spike IgM (blue) at entry, 14d and 28d (n=12).

## Discussion

In this study, we consistently detect SARS-CoV-2 antigen and antibody in oral fluids during symptomatic COVID-19. N-antigen detection was immunoblot- and sgRNA-confirmed in NP-RT-qPCR+ subjects providing significant implications for oral transmission. SARS-CoV-2-targeted oral IgG responses were highly correlated with nasopharynx positivity and oral N-antigen detection. Oral IgM levels indicated both symptom presence and severity.

This study, and others, detected salivary SARS-CoV-2 RNA with distinct viral shedding dynamics compared to NP^1, 21^. Prior assessments detected a 3-fold lower positive detection rate in saliva than NPS^21^, leading some to question the role of oral virus. Here, cumulative data suggest oropharyngeal viral replication. SARS-CoV-2 was detected in oral fluids from NP-RT-qPCR+ participants using several RT-qPCR-based viral detection methods, 1) three distinct primer pairs targeting 3 regions of the viral genome, 2) absolute RNA copy number determination using a standard curve, and 3) sgRNA, shown to be a marker of active replication during early symptomatic infection^22, 23^. Orf3A sgRNA was detected in 82% of those tested by sgRTqPCR, (fast, sensitive, economically feasible, and reliable) and is a viable marker of viral replication based on its contribution to viral titer and disease in hACE+ mice^24^. SARS-CoV-2 protein was consistently detected by two distinct methods, LFA and immunoblot. We are unaware of other studies demonstrating immunoblot confirmed viral antigen detection in oral fluids. Importantly, persistent N-antigen detection provides significant implications for continued potential oral transmission (**Figure 3F,8B**).

One critique of rapid LFA was false positives. Here, we detect positive LFA results in two seropositive (1A,4A), and three uninfected subjects (16A,17A,18A). During asymptomatic infection it is impossible chronical where subjects are in their infection cycle. Seropositive/NP-subjects could demonstrate oral infection (1A,4A) or oral infection may subside in seropositive/NP+ (8A). Alternatively, cross-reactive host proteins might be detected. Analysis determined potential cross-reactivity with structurally analogous host RNA binding proteins (**Figure 5**). 17S U2 snRNP and RBM7, are salivary proteome members and two others, LINE1 Orf1p and hnRNP H, have salivary proteome isoforms (LINE1 Orfp1 and hnRNP A2/B1, hnRNP M, hnRNPK)^25^. This raises the possibility that N-antigen-specific LFA can detect conformationally-similar proteins, perhaps reflecting the potential for N-antigen mimicry through RNA binding domains. An example implication is SARS-CoV-2 host genome integration. The LINE1 ORF1p analogue mediates retrotranposable element genome integration^26^. RNA virus sequences have been detected across vertebrate genomes, with several integration signals consistent with LINE retrotransposon germline integration of viral cDNA copies^27^. Recently, subgenomic sequences, derived from SARS-CoV-2’s 3’ end, were shown to be integrated into host cell DNA^28^. Hence, host interactions with N-antigen or similar actions by N-antigen might facilitate SARS-CoV-2 genome integration.

Anti-spike RBD-specific antibodies during natural infection and vaccination were detected in saliva^29, 30^ with temporal kinetics that reflect blood. While anti-spike-RBD IgG levels were negatively correlated with salivary viral load in the Silva study^31^, here in 16/16 symptomatic and 1/1 asymptomatic participants, anti-spike-RBD IgG was 100% positively correlated with SARS-CoV-2 NP-RT-qPCR+ status, and symptomatic individuals consistently demonstrated salivary N-antigen by immunoblot. The NP+ status and IgG relationship was also shown by Pisanic et,al. with IgG positive responses in 24/24 RT-qPCR-confirmed COVID-19 cases^11^. Together, this suggests that salivary IgG detection indicates concurrent nasopharynx positivity. Detection of simultaneous oral virus and oral host immune responses at baseline suggests a highly infectious pre-symptom state followed by concurrent symptoms, persistent oral infection, and virus-targeted host responses.

Oral fluids hold promise as SARS-CoV-2 prognostic indicators. Huang et al. suggested a correlation between salivary viral RNA burden and COVID-19 symptoms, including taste loss^1^. Others showed positive associations between salivary viral load (VL) and COVID-19-related pro-inflammatory markers; IL-6, IL-18, IL-10, and CXCL10^31^. Here, oral viral RNA detection did not correlate with COVID-19 symptoms. Relationships between IgG, VL, and disease course were not detected^21, 32^. However, we detected statistically significant relationships between salivary IgM and multiple symptoms including weakness, muscle ache, nausea, and upper respiratory symptoms but not taste/smell. Relative IgM levels were significantly associated with degree of fatigue and cough severity suggesting that oral IgM signifies an active early immune response associated with milder disease course.

Gender differences are detected in COVID-19-related morbidity and mortality. Here, gender differences were detected, with women having consistently higher oral IgM levels than men. Systematic review of gender differences determined that women were less likely to present with severe disease and be admitted to the intensive care unit (ICU) than men (OR 0.75 [0.60–0.93] p<0.001 and OR 0.45 [0.40–0.52] p<0.001, respectively^33^. While higher smoking rates and reluctance to seek health care may contribute to male disease, the well-described evolution of greater humoral immunity in females could contribute to these differences^34, 35^. Here, women had markedly higher IgM levels, fewer symptoms, and milder symptom severity. Differences in IgM production may provide biologic underpinning, contributing to the COVID-19-related morbidity and mortality gender gap.

These findings provide novel insights to oral SARS-CoV-2 infection. Oral IgM detection predicted milder disease and female predilection. The potential for cross-detection of N-antigen and structurally similar host RNA binding factors, suggests viral mimicry. Oral persistence has significant implications for viral transmission in the absence of mask use and vaccination.

## Data Availability

All data produced in the present study are available upon reasonable request to the authors

## Acknowledgements

The UNC Delta Core Facility performed sample processing

Biomedomics provide the N-antigen and anti-S-CoV-2 Spike RBD protein IgG/IgM LFA cartridges used in this study.

Institutional Review Board numbers associated with samples collected and used in this manuscript

UNC IRB study #20-0792 – Sample collection from Symptomatic participants

UNC IRB study #20–1771 – Sample collection from Asymptomatic particpants

UNC IRB study # 07-1431 – Sample collection from Archived, PreCovid prticipants

