## Supplemental Material and Methods for "Oral SARS-CoV-2 host responses predict the early COVID-19 disease course"

Enrollment. A total of 47 Covid-19 era participants were assessed. Inclusion criteria for entry into the symptomatic longitudinal observational cohort (n=17) required subjects to be NP SARS-CoV-2 RT-qPCR positive. COVID-19+ patients were recruited after written informed consent and were stratified with mild, moderate or severe symptoms at the time of presentation based on the NIH criteria (NIH COVID-19 treatment guidelines). For the asymptomatic cohort (n=30), participants were either asymptomatic, anti-Sars-CoV-2 spike seronegative (n=15) or asymptomatic, Sars-CoV-2 spike seropositive (n=15) institution campus dwellers including students, staff and faculty who provided written informed consent. Archived, pre-COVID pandemic saliva and throat wash samples were used as SARS-CoV-2 negative controls. All samples were obtained and stored under IRB approval.

Biospecimens For RT-qPCR, NPS, saliva and throat wash samples were collected concurrently. The NPS samples were collected in 3ml of viral transport medium (VTM). Unstimulated whole mouth fluid (WMF) samples (saliva) and throat wash gargle with 10 ml of normal saline (throat wash) were each collected in sterile 50mL wide-mouthed screw-capped containers. Samples were immediately transported for processing where the NPS, saliva, throat wash, and blood samples were stored at 4ºC and processed within 24 hours. After processing 1 ml aliquots of samples were stored at -80°C. Similarly, 1 ml aliquots of archived, preCovid-19 saliva and throat wash samples have been stored at -80°C for >5 years.

Nucleic acid extraction and RT-qPCR. Saliva and throat wash from COVID-19 patients were used as a source for the detection of SARS-CoV-2 RNA. Trizol (Life Technologies, Carlsbad, CA) was used to inactivate virus and RNA was extracted according to the manufacturer’s instructions. Briefly, 750 microliters of Trizol were added to 250 microliters of oral fluid (saliva or throat wash). Following chloroform addition and phase separation, the aqueous phase was collected. Glycogen (2 micrograms) was added to the aqueous phase and RNA was precipitated with isopropanol. RNA pellets were dried and resuspended in water.

RNA was reverse transcribed using Superscript III (Life Technologies) according to the manufacturer’s instructions. First strand synthesis was primed with random hexamers (Life Technologies). First strand synthesis reactions were used in SybrGreen-based qPCR. Three regions of the SARS-CoV-2 genome were targeted using virus-specific primers (**Figure 3A**): WTSorf1F/WTSorf1R, E_Sarbeco_F/E_Sarbeco_R^14^ and 2019-nCoV_N1-F/2019-nCoV_N1-R^15^. A standard curve was generated using SARS-CoV-2 reference RNA (ATCC VR-3276SD) for absolute copy number quantitation.

Subgenomic RNA (sgRNA) detection was performed using the forward primer, CoV25UTR (nucleotides 1-25; GenBank:MN908947) with the reverse primers, SRTqPCRR, orf3aRTqPCRR or 2019-nCoV_N1-R (**Figure 3A**). Percent sgRNA was calculated using the equation: 2^-(sgRNA Ct − N Ct)^ X 100 where sgRNA Ct values were signal obtained using the 5’UTR/specific sgRNA primer pair and the N Ct value was signal obtained using the 2019-nCoV_N1-F/2019-nCoV_N1-R primer pair. Following qPCR, reactions from positive wells were removed, treated with ExoSapIt according to the manufacturer’s instructions (USB, Cleveland, Ohio) and Sanger sequencing was performed (Eton Bioscience, Research Triangle Park, NC) using gene-specific primers: SRTqPCRR for SsgRNA , orf3aRTqPCRR for orf3asgRNA or 2019-nCoV_N1-R for NsgRNA.

Ectopic expression of SARS-CoV-2 proteins in transfected oral keratinocytes. Total RNA was isolated from the saliva of SARS-CoV-2 -infected individuals. RT-qPCR was used to amplify cDNA encoding SARS-CoV-2 Spike (S), Envelope (E) or N-antigen. The generated cDNAs were digest with SfiI and SalI and cloned into SfiI/SalI sites of pCMV-myc eukaryotic expression plasmid. Expression in eukaryotic cells would result in the production of viral proteins with an amino-terminal c-myc tag. Immortalized human oral keratinocytes (NOK) were grown in keratinocyte serum-free media (Life Technologies, Carlsbad, CA). Cells were transfected with expression vectors using Fugene-6 (Promega, Madison, WI). Forty-eight hours post transfection, media was removed and subjected to SDS-PAGE and immunoblot analysis to detect myc-tagged proteins.

To obtain lysates from whole saliva, TW or transfected NOK, protein was isolated from the organic phase obtained using Trizol (Life Technologies, Carlsbad, CA) according to the manufacturer’s instructions. Protein was precipitated after the addition of isopropanol and centrifugation at 12,000 X G. Pellets were washed twice with 95% ethanol/0.3 M guanidine-HCl followed by a final wash with 100% ethanol. Each wash was performed for 20 minutes. Final protein pellets were dried and resuspended in 1X sample buffer (250 mM Tris, 8.0, 2% SDS, 20% glycerol, 50 mM DTT, 0.1% bromophenol blue). Pellets in buffer were incubated at 65° C until dissolved.

Immunoblot analysis of NOK cell lysates and media. Twenty-five micrograms of whole lysate or 10 microliters of transfected cell culture media were loaded onto precast NuPage 4-12% Bis-Tris gels (Life Technologies, Carlsbad, CA). Proteins were electroblotted to PVDF membrane using western blot transfer buffer (25 mM Tris, 192 mM glycine, 20% methanol) for 2 hours at 200 mamps, constant current. Following transfer, blots were blocked by incubation with 5% nonfat dry milk in 1X PBS containing 0.1% Tween-20 (1X PBS-T) at room temperature. Following blocking, blots were incubated with primary antibody overnight at 4° C. Blots were washed twice with 1X PBS-T followed by incubation with HRP-conjugated secondary antibody (Promega) diluted 1:10,000 in PBS-T, 5% milk for 1 hour at room temperature. Blots were washed twice with PBS-T and protein bands were detected by ECL after incubation with ECL Prime (GE Healthcare, Chicago, Il) according to the manufacturer’s instructions. Blots were imaged using a GE ImageQuant LAS4000 system. The following primary antibodies were used to detect proteins on western blots: rabbit anti SARS-CoV-2 nucleocapsid (PA5-114448, Life Technologies), mouse anti-c-myc (sc-40, Santa Cruz Biotechnology, Dallas, TX), anti-amylase (A8273, Sigma-Aldrich, St. Louis, MO).

Overexpression of His-tagged SARS-CoV-2 Nucleocapsid in E. coli. The NcoI/SalI fragent from pCMV-mycN containing the entire coding region of N was removed and inserted into the NcoI/SalI sites of pET30a. The resulting plasmid, containing the SARS-CoV-2 nucleocapsid (N) coding region in frame with the 6X histidine tagged coding region of pET30, was used to transform E. coli BL21 (DE3) bacteria. Bacteria with pET30a empty vector were used as a source of negative control his-tagged protein. Bacteria containing pET30a will produce an ~9 kD his-tagged protein after induction with IPTG. Single colonies were grown in LB medium containing kanamycin (50 micrograms/ml) to an OD A600 of 1.0. To induce gene expression, IPTG (1 mM final concentration) was add and cultures were incubated at 30° C for 2 hours. Following centrifugation of cultures, bacterial pellets were resuspended in 1 ml NPI-10 (300 mM NaCl, 50 mM sodium phosphate, pH8.0, 10 mM imidazole, 1 mg/ml lysozyme). Lysates were incubated with NTA-Ni agarose beads (Life Technologies) to purify his-tagged proteins. Beads were washed with NPI-20 (300 mM NaCl, 50 mM sodium phosphate, pH8.0, 20 mM imidazole). Protein was eluted by resuspending beads in 250 microliters NPI-500 (300 mM NaCl, 50 mM sodium phosphate, pH8.0, 500 mM imidazole). Eluates were subjected to SDS-PAGE followed by Coomassie staining to assess retrieval and protein purity. Protein concentration was determined by Bradford assay.

Lateral Flow Assay. Detection of SARS-CoV-2 N-antigen or anti-SARS-CoV-2 spike protein IgG and IgM was accomplished using lateral flow chambers (BioMedomics, Research Triangle Park, NC). For N-antigen 50 microliters of sample (saliva or throat wash) was diluted with 50 microliter of lysis buffer (supplied by the manufacturer). Samples in lysis buffer were incubated at room temperature. Following incubation, 80 microliters of sample was applied to the lateral flow chamber. Bands were visualized and quantitated using ImageJ software. The control band for each detection cartridge was used to normalize antigen-specific band. For detection of anti-spike IgG/IgM, 50 microliters of sample (saliva or throat wash) was added directly to the lateral flow chamber. Two drops of COVID-19 IgG/IgM rapid test buffer (supplied by manufacturer) were added and bands were allowed to develop. Control bands were used normalize IgG and IgM intensity. Quantitation was determined relative to signal produced by pre-COVID-19 archived saliva and throat wash samples.

NCBI Structural analysis

The crystal structure of the N-terminal RNA binding domain of SARS-CoV-2 N antigen (PDB ID: 6M3M) was compared to other publicly available crystal structures in the Molecular Modeling Database (MMDB) using Vector Alignment Search Tool (Vast+) within the Domains & Structures module on the National Center for Biotechnology Information website (https://www.ncbi.nlm.nih.gov). Vast+ makes geometric structural comparisons between macromolecules in the absence of sequence similarity. The 3D structures of superimposed biological assemblies were visualized using the web-based 3D viewer iCn3D version 4.3.1 to support the visualization style^11^.

Statistical methods

   Given the small sample sizes, non-parametric tests were used to test correlations and compare data. Kendall rank correlation and Mann – Whitney U tests were used to test the correlation between the independent and response variables. The Kendall rank correlation test assessed the existence of monotonic relationships of ordinal or continuous variables using the ranks of the data. The Mann – Whitney U test was used where the independent variables were binary and was used to determine whether the two groups were from the same population when the groups were independent. Mann – Whitney U and Wilcoxon signed rank tests were used to compare response variables between two groups. When the groups were dependent, Wilcoxon signed rank tests were used to assess, differences in the medians of matched groups.
